# Rethinking the Design of Adolescent Crisis Stabilization Units: A Mixed-Methods Study Using Physical Mock-Up Simulations and Artificial Intelligence

**DOI:** 10.64898/2026.01.30.26345181

**Authors:** Roxana Jafarifiroozabadi, Cheng Zhang, Stephen Parker, Virginia Pankey, Hani Patel, Neil Gautam, Chih-Chuan Hsu

## Abstract

Limited research has examined the use of physical mock-ups and artificial intelligence (AI) to evaluate design features in adolescent mental and behavioral health environments, such as the Crisis Stabilization Unit (CSU). This mixed-methods study investigated caregiver workflows and environmental features in adolescent CSUs (e.g., furniture and open vs. enclosed nursing station designs) through physical mock-up simulations with expert and novice clinicians/designers (N = 17). Participants’ feedback was obtained using questionnaires and focus groups. Simulations were video-recorded, manually coded, and an AI-driven tool was developed for automatic analysis of videos. Findings revealed that experts rated the enclosed nursing station higher in visibility, whereas novice designers reported significantly higher perceived privacy in the open nursing station (P = 0.036). AI-driven video analyses demonstrated promising, high-accuracy performance in automatic detecting, tracking, and localizing individuals (>80%) when compared with manual data. This study proposed an innovative methodology to enhance safety in future adolescent CSUs.

## Introduction

Adolescent mental and behavioral health crises continue to rise in the U.S., placing unprecedented demand on care settings designed to provide rapid stabilization and supportive interventions (1,2). Most suspected adolescent mental and behavioral health patients are presented to the general emergency department (ED); however, ED environments are ill-equipped to tackle this situation due to overcrowding, little or no private space for mentally ill patients, long wait times, lack of specialized staff, and environmental hazards (e.g., ligature points) (3–6). A recent report by the Substance Abuse and Mental Health Services Administration (2022) has underscored a critical need to expand non-hospital alternatives to mental health care for young people, such as crisis stabilization units or CSUs (7).

CSU environments have been deemed beneficial in reducing mental health patients’ length of stay in EDs and inpatient facilities (8,9); however, adolescent mental and behavioral health environments still account for high frequency of sentinel events, such as self-harm or aggressive behaviors compromising safety for both patients and caregivers (10). Among the high-risk environments in CSUs is the therapeutic milieus with nursing stations. The milieu environments in CSUs provide a soothing and healing environment for a selected group of patients to de-escalate or receive therapeutic interventions (11). Mental and behavioral care environments such as CSU milieus account for high-risk areas for sentinel events, including assaultive and aggressive behaviors due to environmental factors including poor visibility, space density, understaffing or limited supervision, and organizational factors, such as patient to staff ratios or caregivers working in isolation (10). Despite the critical need in ensuring safety and security in these environments, design features, such as nursing station design typologies and layouts vary widely from enclosed, glass-wrapped nursing stations to open or hybrid designs (12–15). Furniture types and unit layouts also vary significantly, and the existing evidence indicate mixed preferences regarding the optimal design features in mental and behavioral health units (16–18). However, studies to date have mostly focused on adult psychiatric or forensic inpatient facilities and existing resources such as the Facility Guidelines Institute (FGI) or the Department of Veteran Affairs lack specific recommendations or methods to help healthcare designers and stakeholders create safe CSU environments tailored to the specific needs of adolescent patients (19–21). In recent years, physical mock-ups have been used in industry to support, evaluate, and validate decision-making during the design process of healthcare environments for specific patient populations. The Health Quality Council of Alberta (HQCA) has published a framework for conducting simulation-based mock-up evaluations to improve safety in healthcare environments through providing a systematic approach for construction of the mock-up, development of scenarios, evaluations, collecting, and analyzing data (22).

While this framework has been utilized by healthcare design firms and the industry, there has been little attempt to leverage recent advancements in design development and visualization made possible through artificial intelligence (AI) in combination with physical mock-ups. AI-driven monitoring systems can analyze diverse data sources in real-time and thus can offer a unique opportunity for optimizing safety and hazard detection during mock-up simulations. One recent study has even indicated that incorporation of AI-driven tools can predict aggressive or challenging behaviors up to one minute prior to occurrence among youth to improve safety and therefore can potentially play an important role in reducing the rate of sentinel events in mental and behavioral health units (23). Nevertheless, there is still limited knowledge regarding how these tools can be effectively used to analyze care processes and improve safety in adolescent CSU environments. To address these gaps, this study aims to respond to the following research questions: 1) How can environmental design features, such as the nursing station design (open versus enclosed design), furniture type, and layout can impact safety in adolescent CSUs? and 2) How can AI-driven tools be combined with physical mock-up simulations to evaluate care workflows and safety design features in adolescent CSU environments?

## Method

The study used a mixed-methods approach to investigate design features, caregiver workflows, care processes, and safety features in a low-fidelity physical mock-up of an adolescent crisis stabilization unit (CSU) designed and constructed on a university campus in the U.S. The study was conducted in multiple phases: In phase one, a full-scale physical mock-up was constructed and the layout and environmental features of an adolescent CSU, such as furniture and nursing station designs, were investigated with 17 participants using scenario-based simulations (Figure 1). In phase two of the study, the video-recorded simulations were manually coded, and an AI-driven tool was developed to detect, track, and localize individuals in adolescent CSUs based on the recorded simulation videos using computer vision algorithms. The study phases and procedures are described in detail in the following sections.

**Figure 1.**
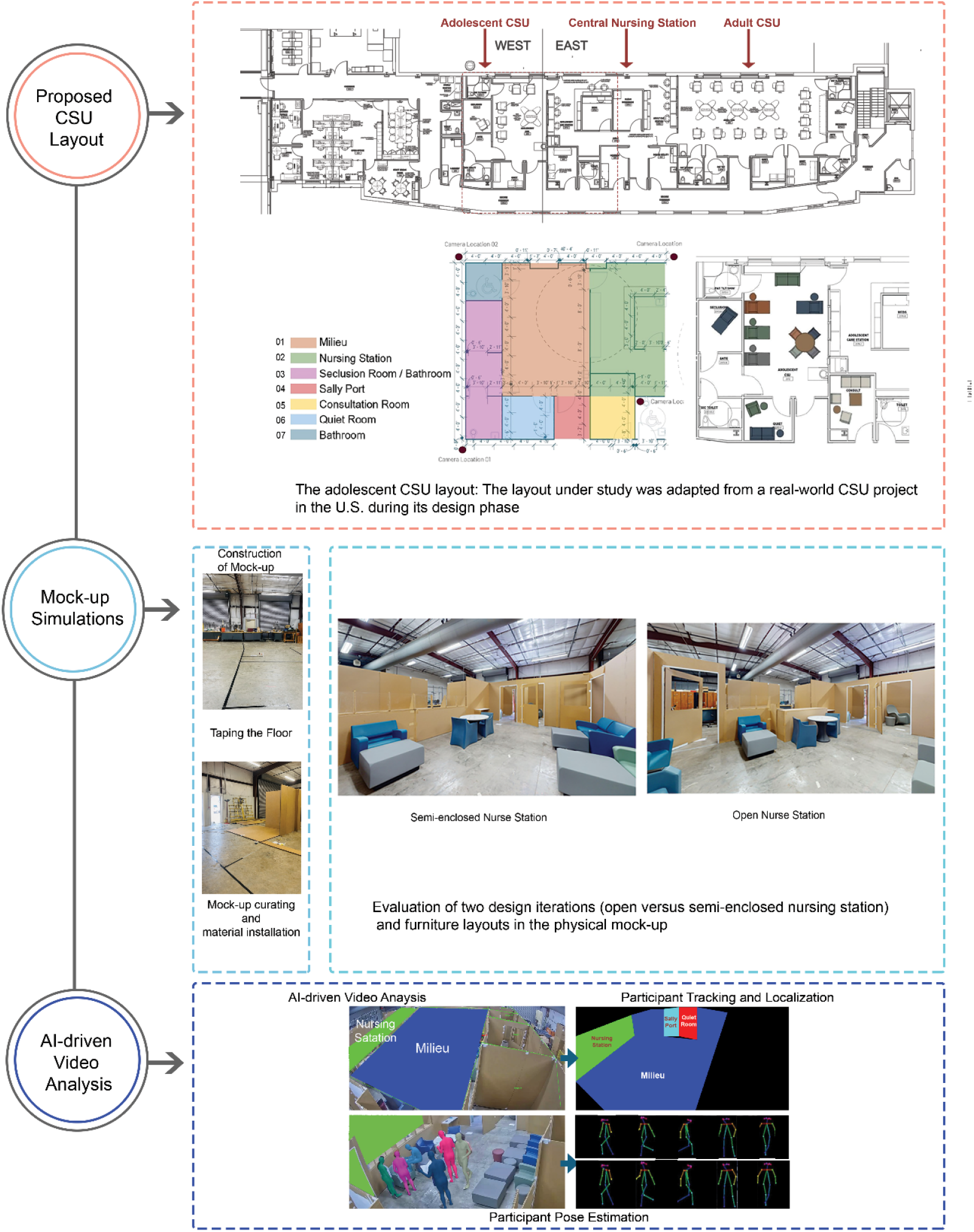
Study procedure conducted in multiple phases. Firstly, the adolescent CSU layout adopted an informed, real-world project in the US. Secondly, a construction of a mock-up and scenario-based simulation took place. Thirdly, AI-driven video analysis occurred.

### Constructing the physical mock-up

The adolescent CSU environment was evaluated through constructing a low-fidelity, full-scale physical mock-up of the unit by a researcher and six research assistants (RAs) on the university campus. The research team evaluated two different design iterations for the nursing station (open versus semi-enclosed design) and furniture layout in the unit within a single lab space during the workshop. The team incorporated the Health Quality Council of Alberta (HQCA, 2020) framework for simulation-based mock-up evaluations of healthcare facilities, which has been adopted by the industry for design evaluation of healthcare environments using scenario-based simulations. The low-fidelity physical mock-up was constructed in four weeks through the following steps: 1) Planning and Design Phase: Concept refinement and taping the floor, 2) Pre-construction Phase: Procurement of materials. 3) Construction Phase: Installation of room features, and 4) Finalization and Evaluation: Detailing, adding surface finishes, final inspections, and documentation.

To facilitate evaluation of multiple iterations, the team constructed modular cardboard panels and installed them in a manner that allowed them to be easily repositioned. The cardboard walls were assembled using custom-made connectors designed by the students to ensure flexibility and ease of reconfiguration, enabling seamless transitions between the design iterations in mere seconds. Following the construction process of the physical mock-up, the team positioned selected types of furniture (e.g., ottomans, sofas, chairs, and recliners) in the CSU milieu for each design iteration based on the recommendations made by a mental and behavioral health furniture manufacturer (Figure 1).

### Scenario-based simulations

The design team designated specific areas within the CSU milieu where participants could role-play to simulate the de-escalation process for an agitated patient with risk of self-harm or harming others. The scenario focused on safety, effective communication, and crisis management in the CSU milieu, incorporating two design iterations for the nursing station and furniture. The scenarios were developed for seven participants with different roles as indicated in the figure 1. The estimated duration of the scenario was approximately 20 minutes.

### Participant recruitment

The expert participants (*N* = 5), including clinicians and behavioral health practice leaders, were selected for participation in the study via purposive sampling across the behavioral health facilities and architecture firms in the U.S. The novice healthcare designers (*N* = 12) were recruited from the College of Architecture on a university campus through purposive sampling using the following inclusion and exclusion criteria: 1) being at least 18 years of age, 2) current or previous recruitment in a graduate-level healthcare design studio, and 3) ability to speak English. The institutional review board (IRB) approval and informed consent were obtained from all participants prior to recruitment and briefing sessions were held to familiarize participants with the study tools and procedures.

## Data Collection and Analytical Strategy

Scenario-based simulations were video recorded for analysis. Data on care workflows participants’ feedback were collected using mixed methods, including investigator-designed questionnaires, focus group discussions, and video-coding of recorded simulations sessions (Table 1).

**Table 1.** Data collection process and instruments.

| **Data Collection Method** | **Variable(s) of Interest** | **Instruments** |
| --- | --- | --- |
| **Investigator-Designed Questionnaire** | Preferences for CSU design features, unit layout, and furniture type | A 16 item-questionnaire related to evaluation of environments safety risks obtained from the Safety Risk Assessment Toolkit for behavioral health units developed by the Center for Health Design (25).<br>A 13-item questionnaire related to evaluation of the furniture type and layout in the unit obtained from the Evidence- Interest Based Design Checklist for healthcare furniture (26). |
| **Focus Group Discussion** | Potential failure modes related to the environment during care delivery | Participants' feedback was recorded using the Failure Modes and Effects Analysis (FMEA) tool during focus group discussions.<br>FMEA is a proactive risk assessment tool endorsed by the Agency for Healthcare Research and Quality, and records scores for Severity (S), Occurrence (O), and Detection (D) of each potential failure mode in the scenario.<br>The parameters are scored as follows: frequency (1 [very low] to 10 [very high]), severity (1, no hazard; 10, extremely hazardous), and detectability (1, almost certain identification; 10, certain nonidentification) (24). |
| **Video- Coding of Recorded Simulation** | Time-based participant tracking and localization | The Noldus observation Labs was used to obtain video and audio recordings of the scenario-based simulations. Four Noldus video cameras were installed in each corner of the mock-up to allow for maximum visual coverage of the care processes. Using the Observer XT software, video-coding was conducted by the team based on the observation of multiple video feeds of simulations simultaneously to determine participant location and roles second-by-second. |

Focus group discussions were audio-recorded and transcribed verbatim for content analysis using MAXQDA (version 2024). Descriptive statistics (means, standard deviations, and percentages) and nonparametric pairwise comparisons were used to analyze focus group and questionnaire data. The FMEA risk priority number (RPN) by multiplying the scores for Severity (S), Occurrence (O), and Detection (D) for possible failure modes during care. High risks are identified when the RPN score is greater than 125 (24). Non-parametric statistical tests were conducted to determine similarities or differences in feedback provided on different design iterations by participants.

To analyze simulation videos, participants’ roles and locations were manually coded second-by-second using the Observer XT (version 17). To automate behavioral mapping during physical mock-up simulations using AI, a video analysis pipeline was developed using computer vision techniques aimed at (1) participant detection and pose estimation (a methodology for locating human body key points (like joints) to understand human posture and actions in images and videos), (2) key participant identification and localization in three-dimensional space in real time in videos. The performance accuracy of the video analysis pipeline was investigated through pairwise comparisons of second-by-second participant detection rates with manually coded video data. To analyze the videos, the simulation videos were cut into 180-second clips, and each clip into 30-second clips (30 fps). Each frame of the simulation videos was processed using a state-of-the-art object detection model (YOLO), to identify the presence and location of people in the scenes (27). Participants in simulation videos were manually annotated with bounding boxes and assigned unique identifiers, enabling consistent tracking throughout each simulation scenario. All statistical analyses were conducted using RStudio (RStudio Team, 2023).

## Results

### Participants

Experts (N = 5) consisted of behavioral health practice leaders (n = 3) and clinicians (n = 2), aged 30–60 years, predominantly female (n = 4), and holding master’s or doctoral degrees. Novice healthcare designers (N = 12) were graduate-level healthcare design students at the university, all with bachelor’s degrees, aged 18–30 years, predominantly female (n = 7) and White/Caucasian (n = 8).

### Questionnaire Data

Experts rated the semi-enclosed nursing station higher than the open nursing station in terms of visibility (M = 4.2, SD = 0.45 vs. M = 2.8, SD = 0.84) and elimination of blind spots (M = 3.9, SD = 0.89 vs. M = 2.2, SD = 1.09). Experts rated visual and acoustic privacy slightly higher in semi-enclosed nursing stations compared to open ones (M = 3.67, SD = 0.82 vs. M = 3.20, SD = 0.84). Novice healthcare designers rated the open nursing station design slightly higher in terms of caregivers’ visibility compared to the semi-enclosed design (M = 3.67, SD = 1.03 vs. M = 3.33, SD = 0.82). Comparing ratings between experts and novice designers using the Wilcoxon rank-sum test (Mann–Whitney U) indicated that novice designers ranked perception of privacy significantly higher than experts for the open nursing station design at α =.05 (Z = –2.10, p = 0.036). No statistically significant differences were observed between experts and novice designers’ ratings of other variables.

While there were discrepancies in participants’ feedback regarding nursing station designs, both experts and novice healthcare designers unanimously evaluated stability (M = 5.00, SD = 0.0 vs. M = 4.73, SD = 0.47), ligature resistance (M = 4.80, SD = 0.45 vs. M = 4.64, SD = 0.50),, and edge safety (M = 4.60, SD = 0.89 vs. M = 4.73, SD = 0.47) highly in the unit. However, novice designers rated the seating’s ability to support food-positioning changes significantly lower than experts at α =.05 (Z = 2.29, p = 0.011) in the physical mock-up simulations. Figure 2 illustrates the summary of results from the questionnaires.

**Figure 2.**
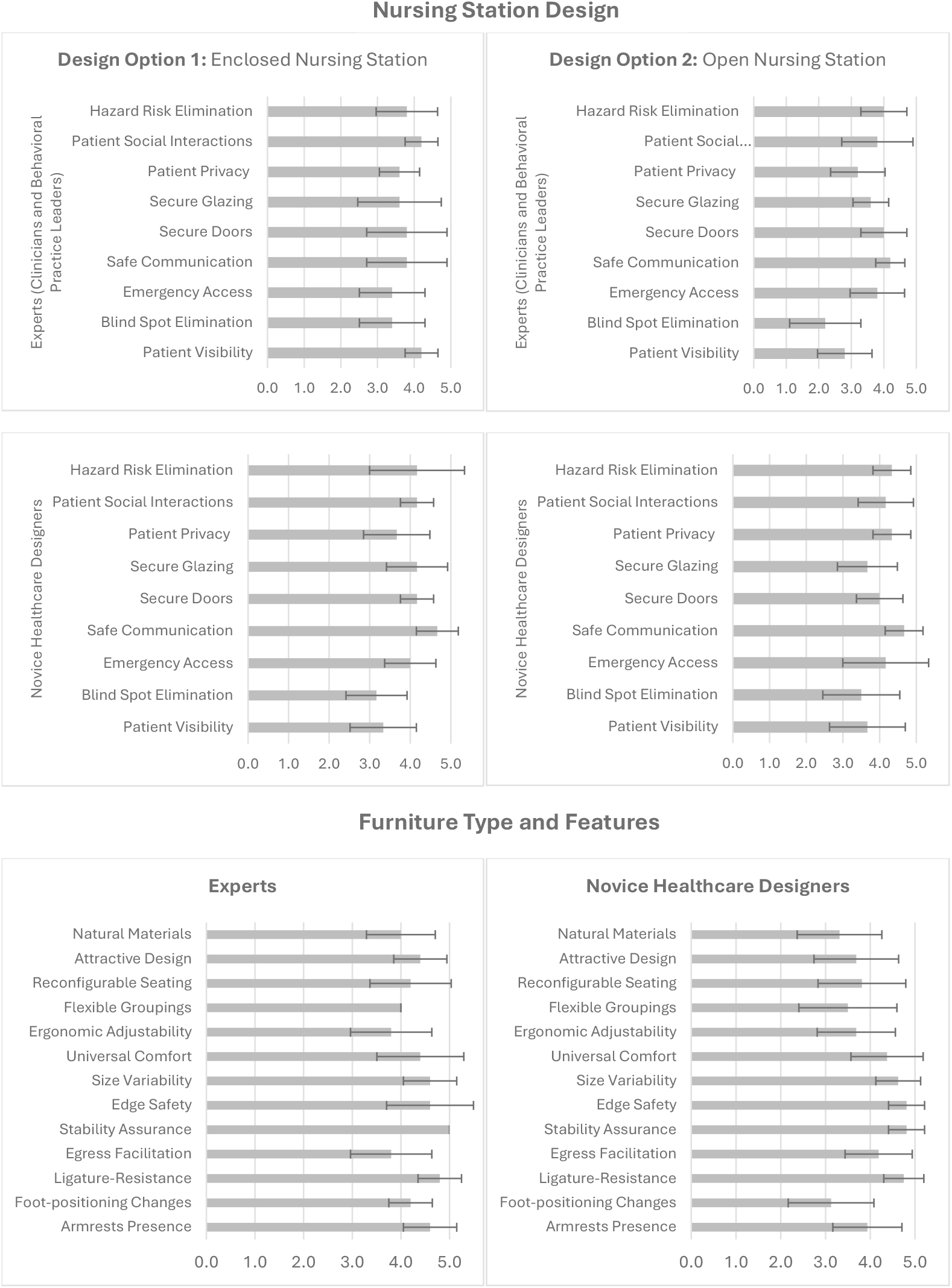
Summary of questionnaire results comparing nursing station design and furniture feature evaluations in the adolescent CSU mock-up between experts and novice designers.

### Focus Group Discussions

The results were analyzed by the FMEA tool indicated that experts and novice healthcare designers identified cluttered furniture layout close to the nursing station as a failure mode with average RPNs of 500 and 640, respectively. Experts also identified blind spots in the milieu environment with open nursing station design (average RPN = 500), and proposed design strategies, such as round corners or including a charting station in the milieu to improve sightlines and visibility in the unit. In addition, both experts and novice healthcare designers proposed incorporating secondary doors adjacent to the open nursing station, enabling controlled access to the milieu during emergencies with average RPNs of 1000 and 850, respectively. Moreover, participants proposed alternative placements for the seclusion zone due to lack of visibility and accessibility for caregivers from the nursing station. Table 2 illustrates the summary of FMEA tool assessments by experts versus novice healthcare designers and the RPN score associated with each failure mode identified during discussions.

**Table 2.**
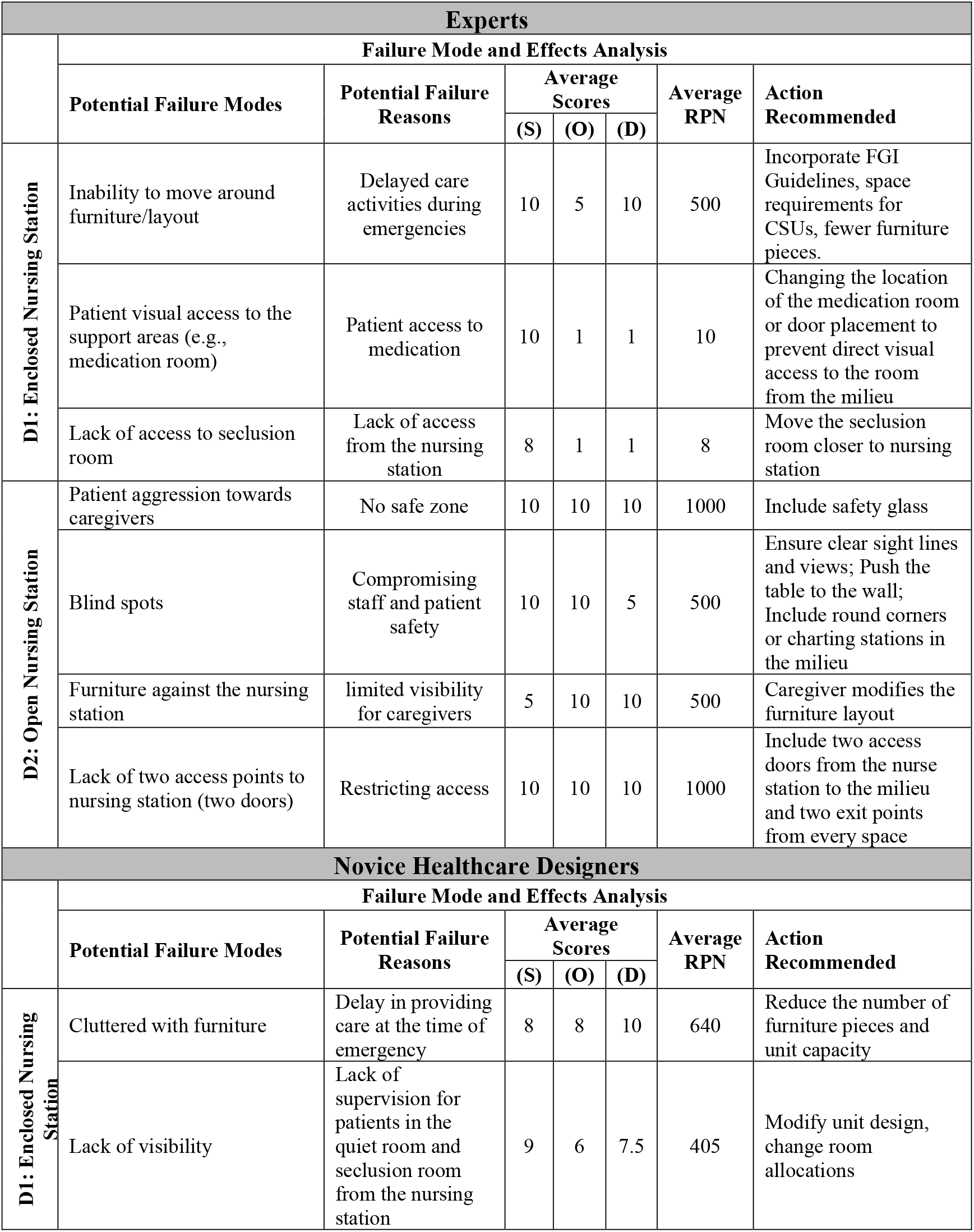
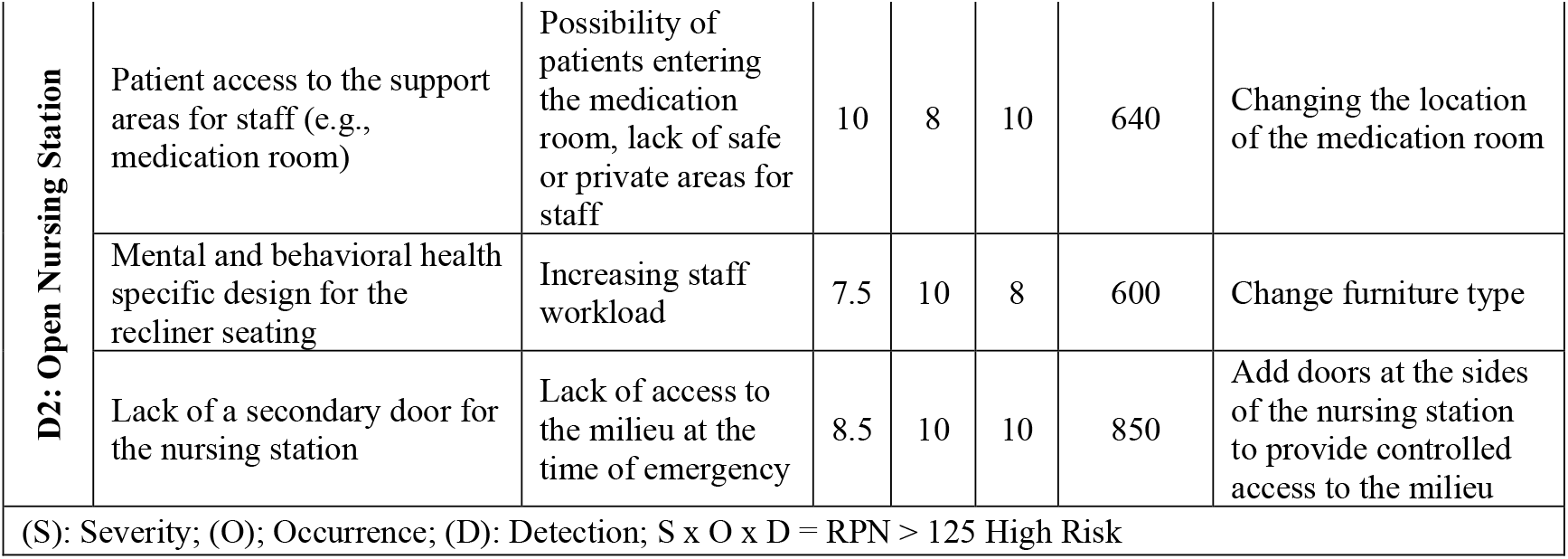
Summary of the focus group discussions.

### Enhancing the Performance of the Video Analysis Pipeline

Following manual annotation of videos with bounding boxes and assigning unique identifiers for tracking participants using YOLO, the performance of the tool was evaluated using second-wise comparisons against the ground truth (GT) data generated through manual coding of videos (Figure 2). Nevertheless, initial evaluations of the tool indicated that detection accuracy, defined as the number of correctly predicted seconds divided by the total number of annotated seconds, fell below 80% for some participants due to occlusion in the video frame and participant movements. To enhance the tool performance, a tracking algorithm (DeepSORT) was used to assign persistent labels to each detected participant based on appearance and motion cues across time. To preserve participant anonymity during analysis, the Human4D model (28) was applied to estimate the 3D body pose and shape of each participant by color-coded meshes overlaid on the original video, fully substituting visual textures with a simulated 3D representation. To improve participant identification, a pre-trained model (ResNet50) was used to recognize visual features from the video frames (29). A feature bank was also created using 40 sample images of each participant, along with generated features to cover different appearances and improve recognition accuracy across various video angles. For each frame of the video, the pose estimation model (Human4D) produced a set of key points (e.g., skeletal joints) for each participant. To determine which detections in the current frame corresponded to the same individuals in the previous frame, the system calculated the L2 distance (Euclidean distance) between key point coordinates of every possible pair of detections and produced a cost matrix representing the spatial similarity between detections across frames. The Hungarian Algorithm was then applied to the cost matrix to find the optimal one-to-one assignment of detections between frames, minimizing the total matching cost. The improved accuracy of automated participant detection, tracking, and localization across the videos is indicated in Figure 3.

**Figure 3.**
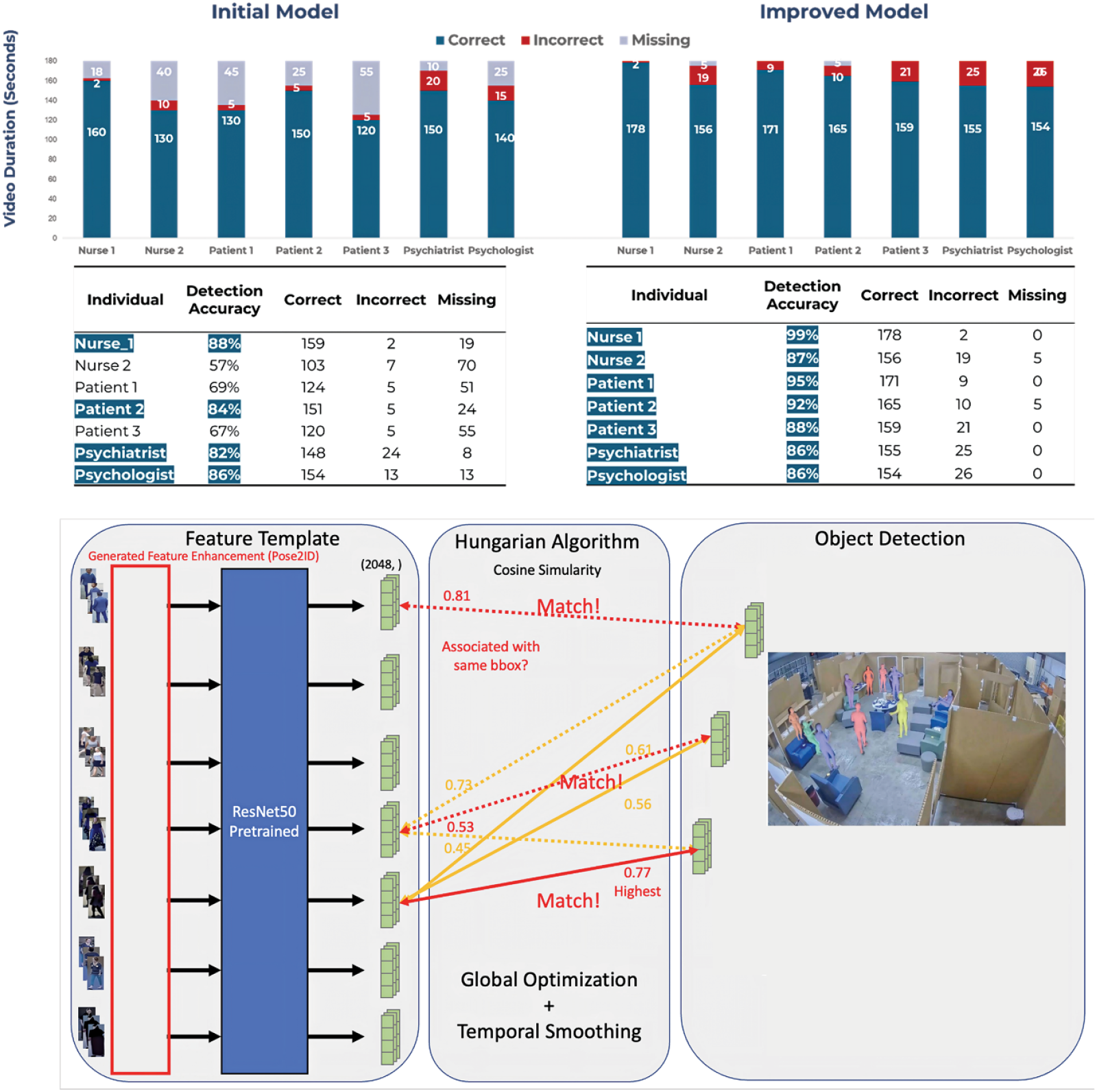
The performance improvement process of the AI-driven video analysis pipeline.

## Discussion

This study used mixed-methods incorporating scenario-based simulations in a physical mock-up and AI-based video analysis to evaluate environmental design features (e.g., nursing station design, furniture type, and layout), care workflows, and their impact on safety in adolescent CSU environments. Findings indicated discrepancies in the evaluation of design features, such as open versus semi-enclosed nursing stations in the CSU between experts and novice designers participating in scenario-based simulations. While enclosed nursing station accounted for higher ratings for visibility and lack of blind spots among experts (clinicians and behavioral practice leaders), novice designers ranked those variables higher for open nursing stations in the CSU. These findings are in line with mixed preferences for open versus enclosed nursing stations reported in existing studies. While some studies emphasized how enclosed nursing stations support safety, privacy, and confidentiality of patients’ health information in therapeutic milieus (12,15), others have revealed that open nursing station designs in milieu environments were commonly associated with enhanced patient-staff communication and perception of a home-like feeling in units (30,31). Although some studies have promoted open nursing station designs in milieus to enhance patient-centeredness, they still recognize significant safety and physical security concerns, especially regarding access to potentially dangerous items in open stations (14,32). Moreover, most of these studies to date have focused on the adult mental and behavioral health units, such as inpatient psychiatric units or forensic units, and have not investigated the preference of nursing station designs in CSUs accommodating adolescent patients.

Focus group discussions also emphasized the importance of visibility, access, and spatial density based on high RPN scores among participants and revealed the importance of nursing station location in the adolescent milieu and its adjacency to certain areas such as the seclusion suite. Existing studies have also emphasized the importance of proximity to key patient areas such as for maintaining oversight and safety, irrespective of the nursing station’s design, while highlighting that enclosed nursing stations could optimize observation capabilities and improved sense of control among caregivers (17,33). Despite discrepancies in participants’ feedback regarding the nursing station design in the adolescent milieu environment, all participants unanimously agreed on most important furniture features, including stability, ligature resistance, and edge safety. Lack of such features have also been noted in existing studies as potential safety risks with furniture being used as weapons or to hide contraband in mental and behavioral health units (17,34).

Exploring the incorporation of AI in analysis of simulation videos for detecting, tracking and localizing individuals in the CSU milieu environment showed promising results with high levels of accuracy when compared with manually coded data. Validating the data obtained from the developed video pipeline against manually coded data helped improve the performance and accuracy of the AI-driven tool to automatically detect and track participants in the unit. However, there is still limited understanding of how AI can be applied to enhance safety or acceptability in youth mental and behavioral health environments, and important ethical and privacy-related concerns continue to surround the use of this technology. While application of the Human4D model for 3D body pose estimation facilitated anonymous tracking of participants, additional research is required for consistent anonymity during real-time observations and tracking of individuals considering that the present study primarily focused on retrospective analysis of videos using AI-driven tools. Other limitations of this study include: 1) this study was conducted in the lab setting therefore generalizing findings of this study might not be applicable to other facility types or the real-world setting, 2) the experts who joined this study were mainly behavioral practice leaders and psychologists and additional studies are needed with patient-caregiver dyads to reflect on optimal and safe design features in adolescent CSU environments. Future studies can also evaluate the proposed layout with actual adolescent patients to obtain their feedback regarding the environmental features in CSUs.

## Conclusion

This study contributes to a growing body of research on behavioral health facility design by focusing on crisis stabilization environments specifically tailored for mental and behavioral health adolescent patients. By using an innovative approach combining physical mock-up simulations and AI, the study identified safety, environmental design features (e.g., the nursing station, furniture type, and layout) to enhance safety in adolescent CSU environments; however, findings revealed a notable gap in preferences of design features in adolescent CSUs between participants involved in the design process of such environments. The study also explored AI application for improving safety through tracking individuals and analysis of clinical workflows in CSU environments. While findings showed promising results in accurate identifying and tracking individuals, additional research is required for incorporation of AI in analysis of behavioral patterns and activities in units to mitigate the risk of sentinel events in adolescent CSU environments.

## Data Availability

All data produced in the present study are available upon reasonable request to the authors

## Acknowledgement

The authors would like to thank the Texas A&M College of Architecture, Norix, and Solace furniture for their support for this study.

## Notes

**Declaration of Conflicting Interests** The Authors declare that there is no conflict of interest.

**Funding** This work was supported by The College of Architecture at Texas A&M University.

### Competing Interest Statement

The authors have declared no competing interest.

### Funding Statement

This work was supported by The College of Architecture at Texas A&M University.

### Author Declarations

Ethical approval for this study was obtained from The Texas A&M University Institutional Review Board, STUDY2024-0920. All participants provided written consent before they participated in the study.

